# The Causal Effect of Air Pollution on COVID-19 Transmission: Evidence from China

**DOI:** 10.1101/2020.10.19.20215236

**Authors:** Guojun He, Yuhang Pan, Takanao Tanaka

## Abstract

There is increasing concern that ambient air pollution could exacerbate COVID-19 transmission. However, estimating the relationship is challenging because it requires one to account for epidemiological characteristics, to isolate the impact of air pollution from potential confounders, and to capture the dynamic impact. We propose a new econometric framework to address these challenges: we rely on the epidemiological Susceptible-Infectious-Recovered-Deceased (SIRD) model to construct the outcome of interest, the Instrument Variable (IV) model to estimate the causal relationship, and the Flexible-Distributed-Lag (FDL) model to understand the dynamics. Using data covering all prefectural Chinese cities, we find that a 10-point (14.3%) increase in the Air Quality Index would lead to a 2.80 percentage point increase in the daily COVID-19 growth rate with 2 to 13 days of delay (0.14 ∼ 0.22 increase in the reproduction number: R_0_). These results imply that improving air quality can be a powerful tool to contain the spread of COVID-19.

## Introduction

The rapid spread of COVID-19 has become a massive global public health and economic crisis, bringing millions of infections^1^ and massive layoffs^2^. To design an effective response to this unprecedented pandemic, it is important to identify factors affecting virus transmission. Existing studies have documented that age^3,4^, gender^4,5^, comorbidities^3,5,6^, and climatic conditions^7^ affect the virus symptoms. However, less is known about how ambient air pollution, which causes severe damage on various health outcomes^8^, can affect virus transmission.

Ambient air pollution could affect the spread of infections through increasing both exposure and susceptibility to the virus (see Supplementary Note 1 for details). Air pollution can cause a persistent inflammatory response and impair the respiratory and immune systems^9,10^, making it more challenging for an individual to resist infection. Besides, recent studies suggest that aerosols in the air may maintain the viability and transmissibility of the virus^11-13^. Therefore, degraded air quality, dominated by particulate matter (i.e., aerosols), may extend the survival of the virus in the air, which amplifies the chances of infection. Several previous papers suggest that air pollution may increase the spread of influenza^14,15^ and SARS^16^; more recently, it has been found that air pollution is correlated with COVID-19 incidence in the U.S.^17,18^, Germany^19^, the Netherlands^20^, Italy^21^, the U.K.^22^, and China^23^.

However, at least two limitations have plagued the existing studies linking ambient air pollution to the COVID-19 outbreak. First, unlike other health outcomes, the virus growth is exponential, and failure to account for such non-linearity could easily provide biased estimates. For example, most existing studies use linear regression models to quantify the relationship between air pollution and COVID-19 prevalence and use new COVID-19 confirmed cases or deaths as the outcomes^17-23^. However, such models do not consider the epidemiological features of disease transmission and may mistakenly generate spurious correlations between air pollution and factors that could affect the virus spread. (In the Methods section, we describe in detail why this is the case).

Second, quantifying the causal effect of air pollution on COVID-19 transmission is challenging. It is well documented in the literature that omitted variables and measurement errors could generate substantial biases in estimating the air pollution effects^24-31^. Such concerns are exacerbated in the case of COVID-19, because economic activities (e.g., opening industries and schools), health interventions (e.g., social distancing and business closure), and avoidance behaviors (e.g., wearing masks) not only change the transmission of COVID-19, but also could be correlated with air pollution exposure^32,33^. Existing evidence on the relationship between air pollution and COVID-19 relies mostly on associational approaches to quantify the impact and does not explicitly address the endogeneity of air pollution levels^17,19,21-23^. Hence, it remains unclear to policymakers, healthcare professionals, and researchers whether air pollution can causally affect the transmission of COVID-19.

In this study, we estimate the plausibly causal effect of air pollution on the transmission of COVID-19 across China, where the levels of air pollution are orders of magnitude higher than those in developed countries. Our empirical strategy is designed to overcome the two challenges. First, our econometric specification is derived from the Susceptible-Infectious-Recovered-Deceased (SIRD) model, which is widely used by epidemiologists to characterize the transmission of infectious disease. Based on this model, we show that the use of daily confirmed cases (or deaths) as the outcome variable in the regressions, as commonly employed by previous studies, could be problematic^17-23^ (see Methods for details). Instead, the daily growth rate of the confirmed active infections is used as the outcome variable; it is obtained by taking the first difference in the natural logarithm of daily confirmed active cases. The model implies that using growth rate as the outcome variable allows us to account for the exponential epidemic growth.

To address the second challenge, we adopt an Instrumental Variable (IV) approach to isolate the impact of air pollution from potential confounders. Specifically, we employ thermal inversions to instrument air pollution. Thermal inversion is a natural phenomenon that involves changes in the normal tendency of the air to cool down with altitude. When a thermal inversion occurs, a layer of warmer air overlays a layer of cooler air in the atmosphere^25,34^. Because warmer air has a lower density, the air pollutants emitted from the ground surface are “trapped,” which eventually leads to higher levels of local air pollution. The intuition of the IV model is that, while observed air pollution could be correlated with local economic activities, health interventions, individuals’ averting behaviors, etc., part of the variation in air pollution can be exogenously changed by thermal inversions. Thermal inversion is a complicated meteorological phenomenon and is unlikely to be correlated with the confounders mentioned above (see Methods). Therefore, by exploiting changes in air pollution that are induced by the plausibly random occurrences of thermal inversions, we can credibly isolate the causal impact of air pollution from the confounding factors.

In addition, to examine the dynamic impact of air pollution on the transmission of COVID-19, we incorporate the Flexible Distributed-Lag (FDL) models into our IV estimation^38^. Accounting for its dynamic component is important because there are time delays between infections and case confirmations due to the period of incubation, testing, and reporting^35-37^. Moreover, to ensure that our regression controls for potential confounders, we include date fixed effects to account for shocks identical to all cities but unique to each date (such as macroeconomic condition, the national level change in the definition of the disease, and the nation’s virus containment policies) and city fixed effects to account for time-invariant characteristics unique to each city (such as local healthcare resources, local testing capacity, and short-term population structure). We also include time-varying weather variables (temperature, precipitation, and snow depth) in the regressions.

The core of our analysis uses comprehensive COVID-19 incidence and air quality data at a day-by-city level from 1 January to 1 April 2020 (N=30,360) in China (Figure 1). We focus on China because COVID-19 was largely controlled in the country by 1 April 2020, so we can observe the entire cycle of the virus transmission. The air quality data are collected from 1,605 monitoring stations covering all the 330 prefectural cities. We focus on the Air Quality Index (AQI), a composite measure of air pollution adopted by the Chinese government (see Supplementary Figure 1 and Supplementary Table 1 for details). The daily number of confirmed COVID-19 cases in each city is obtained from the National Health Commission of China. There were 49,982 confirmed cases in Wuhan city and 30,441 cases in other cities by 1 April 2020 (Figure 2 and Supplementary Figure 2). In our baseline analysis, we exclude Wuhan city from the regressions because of concerns about the city’s COVID-19 data quality^39^, but our results remain robust when we include data from Wuhan.

**Figure 1.**
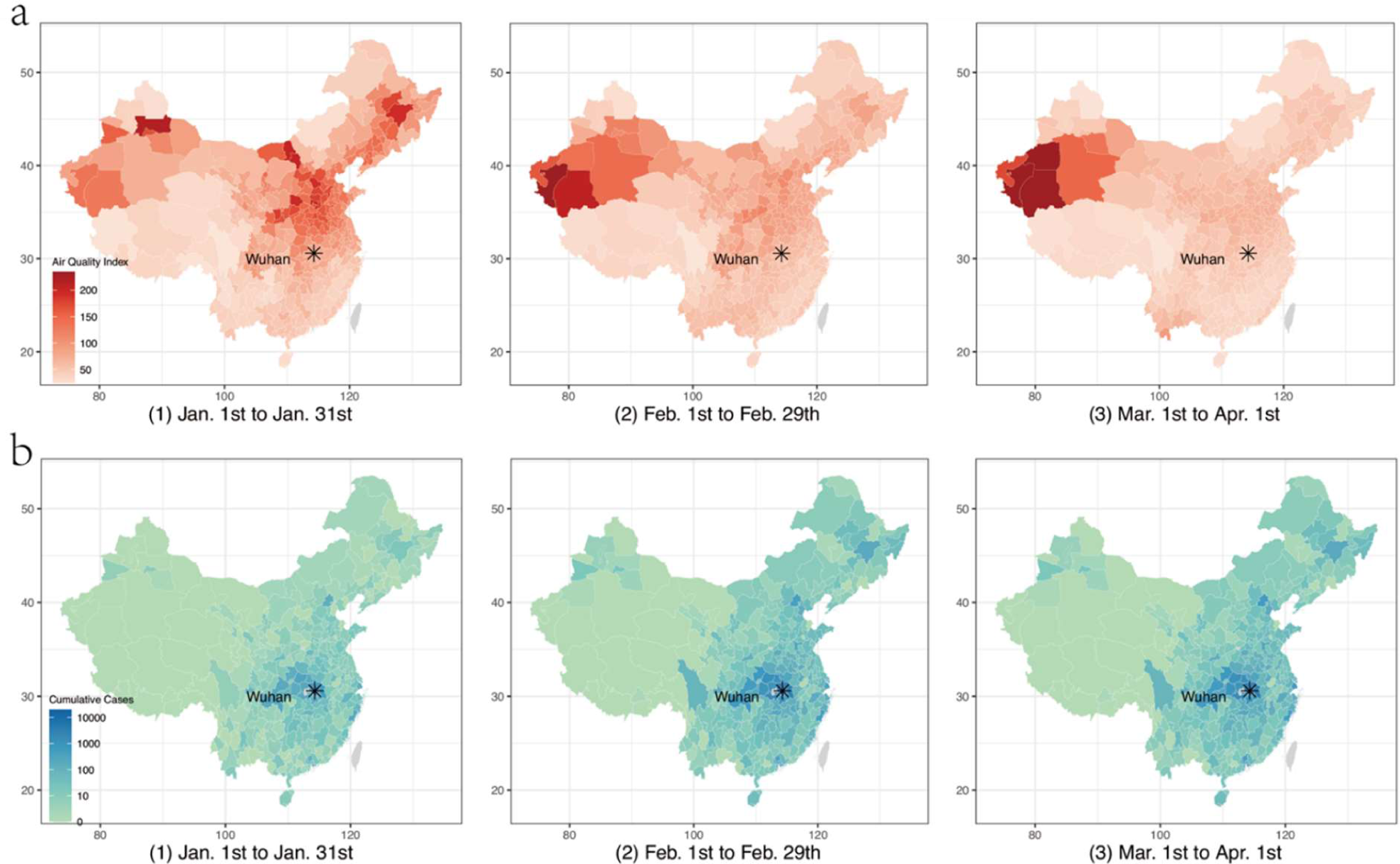
Data on COVID-19 infections and air pollution in China. **A**. The trend of the Air Quality Index (AQI). Higher AQI means worse air pollution. AQI is a comprehensive measure of air pollution: the index is constructed using PM_2.5_, PM_10_, SO_2_, CO, O_3_, and NO_2_ concentrations (See Methods Materials). **B**. The Confirmed COVID-19 cases. The gray color denotes the no-data area.

**Figure 2.**
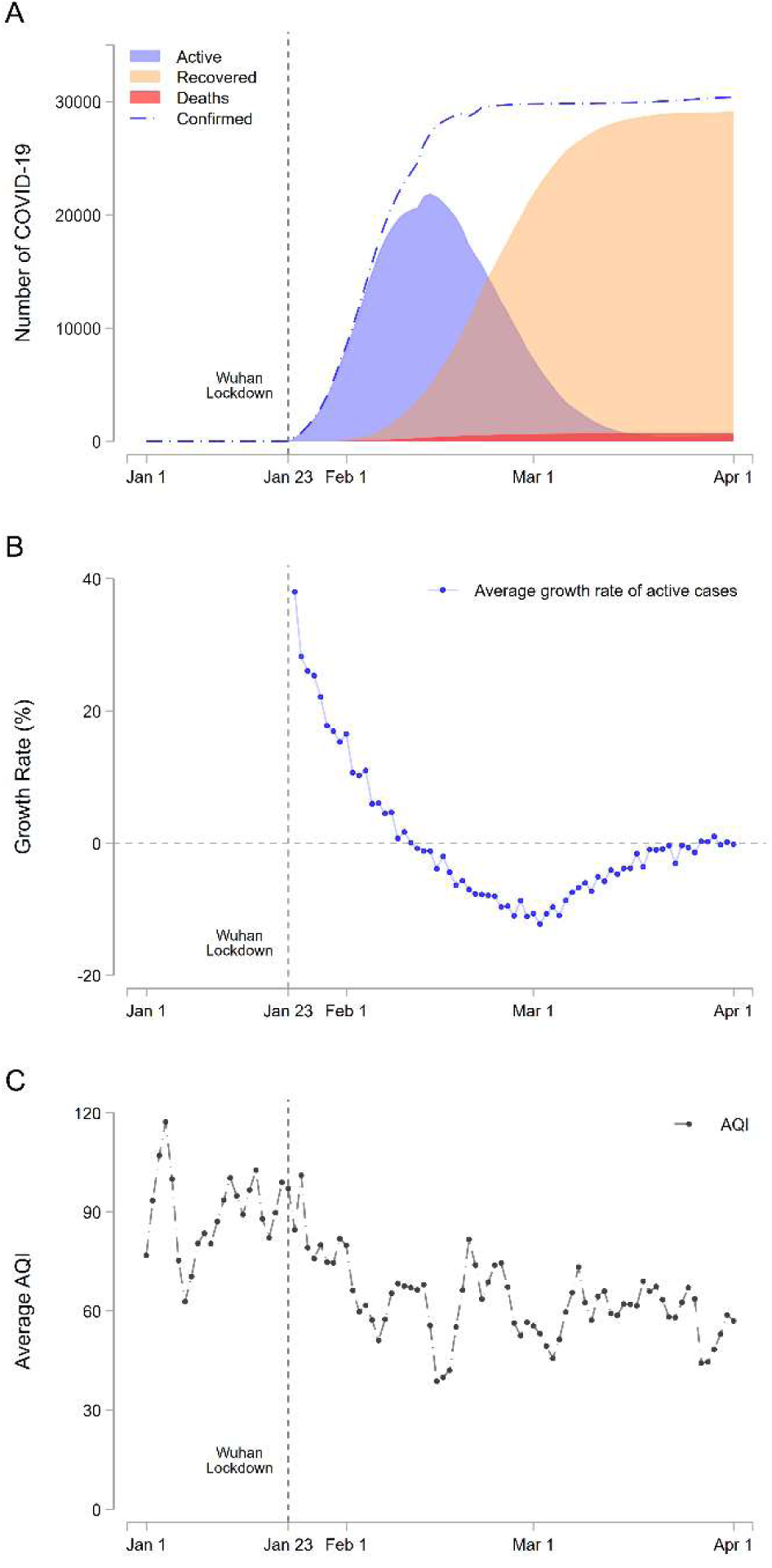
Data on COVID-19 infections and air pollution. **A**. Daily confirmed cases (blue dash line), active infections (blue area), recovered (orange area), and deaths (red area) from 1 January to 1 April. The vertical line is the date of the Wuhan lockdown (23 January). **B**. The average growth rate of active infections. If it is larger than 0, the active infections increase. **C**. The trend of the Air Quality Index (AQI). Higher AQI means worse air pollution. AQI is a comprehensive measure of air pollution; the index is constructed using PM_2.5_, PM_10_, SO_2_, CO, O_3_, and NO_2_ concentrations (See Methods Materials).

## Results

The IV estimates can be obtained through a Two-Stage-Least-Squares (2SLS) procedure. In the first stage, we predict the air pollution concentration using thermal inversion (temperature difference between the ground surface and the upper layer). In the second stage, we regress the daily virus growth rate on the predicted pollution concentration rather than on observed pollution concentration. While observed air pollution could be correlated with factors potentially affecting the virus prevalence (economic activities, health intervention, and averting behaviors), air pollution concentration derived from the thermal inversion is plausibly uncorrelated with such confounders.

### First-stage: Thermal inversion and air quality

For the 2SLS procedure to be valid, it has to be the case that the thermal inversion is a strong predictor of the air pollution concentration. We find that thermal inversion is strongly correlated with variation in local air pollution, even when conditioning on all the time-varying control variables and a set of fixed effects (Figure 3A). Specifically, a 1°C increase in the temperature inversion is associated with a 3.03 point increase in the Air Quality Index (AQI) (Figure 3B) – where a higher AQI means worse air quality. Such patterns are also found when we further control for the city lockdown indicator and the days since the disease outbreak, and when we include Wuhan in the regression (Supplementary Figure 3). To confirm that this strong relationship is not driven by the local time trends or the spatial distribution of frequency of thermal inversions, we randomly shuffle the observed thermal inversions within the same location or within the same day 1000 times. The average estimates using the placebo sample in each case are close to zero, implying that the relationship is not spurious (Figure 3B).

**Figure 3.**
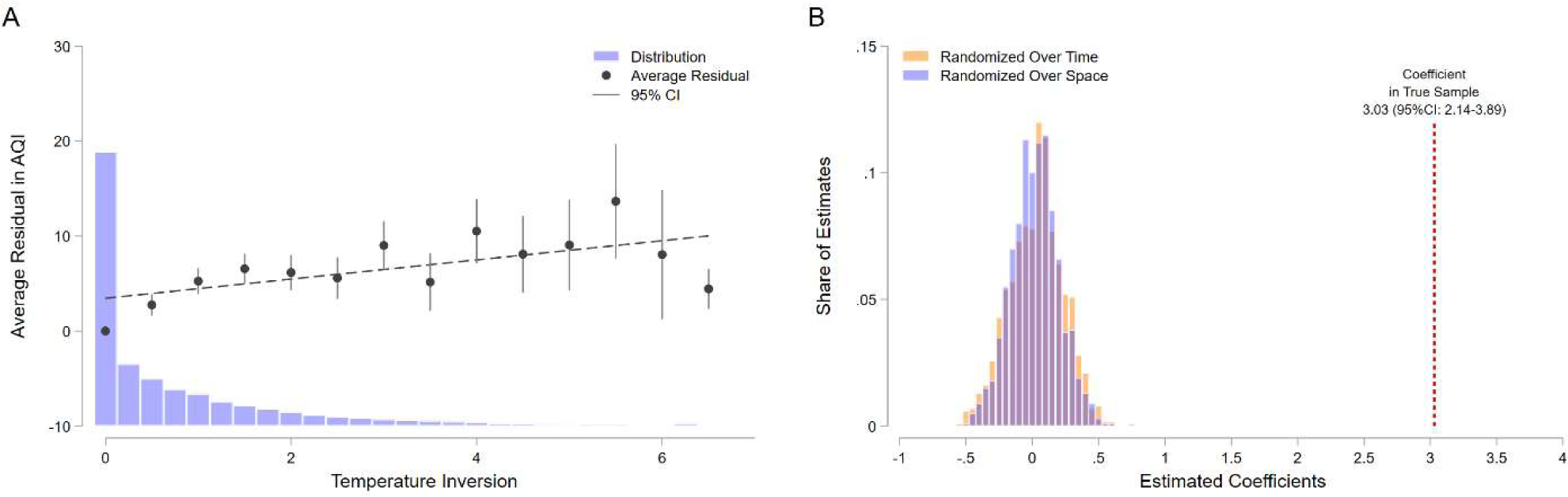
Variation in thermal inversion is strongly correlated with the Air Quality Index. **A**. The graph represents the distribution of thermal inversions, defined as the difference between the temperature in the second layer (320m) and that in the first layer (110m). The Y-axis represents the residual in variation in the AQI after controlling for temperature, precipitation, snow depth, city fixed effects, and date fixed effects. A higher temperature in the second layer is strongly associated with a larger residual in the AQI (worse air quality). **B**. A 1°C increase in temperature inversion is associated with a 3.03 increase in the AQI. In contrast, we find no statistically significant association between placebo thermal inversion and the AQI. In the placebo sample, we randomized the thermal inversions across different days within the city, or across different cities within a specific day.

### Second-stage: Dynamic relationship between air quality and COVID-19 growth rate

In the second stage, we use predicted air pollution from the first-stage regression to estimate the pollution-transmission relationship. We include up to 22 days of lags in the regressions (current + previous 21 days) to capture its dynamic impacts.

Our second-stage regression demonstrates positive impacts of air pollution on the COVID-19 growth rates with 2 to 13 days of delay (Figure 4A, see Supplementary Table 2 for full results). This delay is consistent with epidemiological observations that the disease is usually confirmed after incubation, testing, and reporting^35-37^. Specifically, a 10 point (14.3%) increase in AQI during these windows raises the growth rate by 2.80 percentage points. In contrast, before and after these periods of higher pollution, air pollution does not have a consistent and meaningful effect: if we add the impact before t-2 and after t-14, the size of joint coefficients varies only slightly, but its standard error becomes larger.

**Figure 4.**
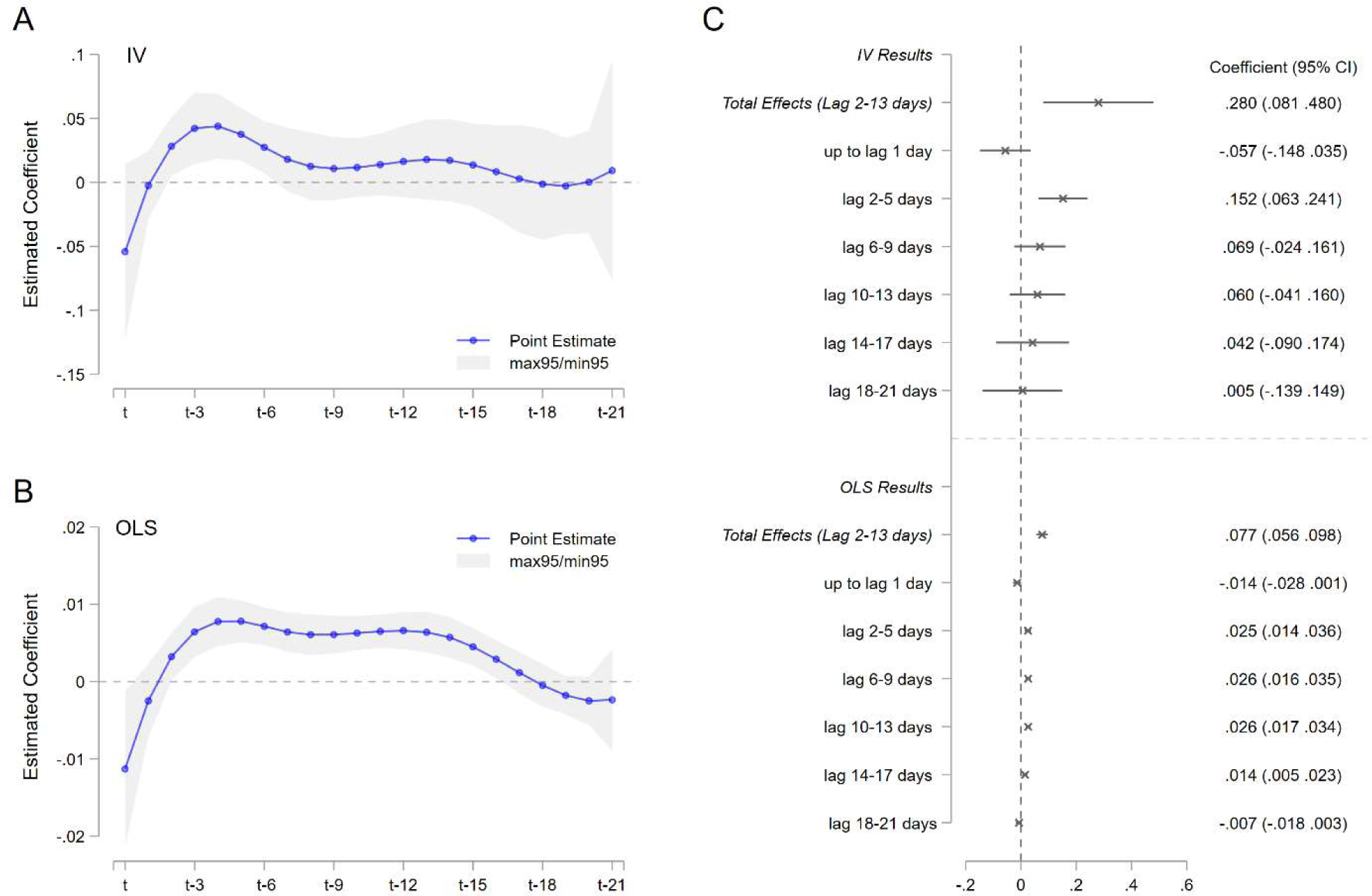
Severe air pollution amplifies COVID-19 transmissivity with 2-13 days of delay. **A**. shows the results using 2SLS. Weather controls (temperature, precipitation, and snow depth), date fixed effects, and city fixed effects are included in both the first and second stage regression. The blue line represents the point estimates, while the gray area denotes the 95% confidence interval. **B**. directly regresses the daily disease growth rate on the observed air pollution rather than predicted air pollution. The regression includes the same controls as IV estimates. **C**. compares the magnitude of the effects between IV results and OLS results. The gray crosses represent the point estimates, and the gray line is the 95% confidence interval. In all regression, standard errors are clustered at the city level.

These results are robust to a number of different model specifications. We add the city’s lockdown status (Supplementary Figure 4A) and days since the outbreak as control variables (4B), include Wuhan in our regression (4C), and use different FDL model segments settings (4D, 4E). All results remain similar. In addition, we add three days of future air pollution as another placebo test and find that, as expected, its effects are close to zero and statistically insignificant (4F). When we change the lengths of the lags of air pollution from 16 days to 24 days, we also observe similar patterns (Supplementary Figure 5). (see Supplementary Note 2 and Supplementary Table 3 for the city’s lockdown data^40^).

We observe that the average daily growth rate in the first week of the epidemic outbreak is 24.9% across cities, with the doubling time of infections at 2.78 days. If AQI increases 10 points, the doubling time would be shortened to 2.50 days. Alternatively, assuming the removal rate (total of recovered rate (*γ*) and death rate (*ρ*) in our SIRD model) is 13%∼20%, which means the patients recover or die in 5.1∼7.1 days (see Supplementary Note 3), a 10-point increase in AQI would lead to 0.14∼0.22 higher reproduction number (*R*_0_ = *β*/(*γ* + *ρ*)).

Existing studies linking air pollution to various health outcomes often find that ordinary least squares (OLS) estimates understate the impact of air pollution^24-31^ (Supplementary Figure 6), in that observed air pollution is directly used. We also find that the OLS estimates are substantially smaller than the IV estimates (even though the dynamic patterns are similar). OLS regression shows that a 10-point increase in AQI between 2 days and 13 days before the case is reported is associated with a 0.77 percentage point increase in the growth rate. This is about one-fourth of the IV estimate, suggesting that the OLS estimate can be biased downward. Supplementary Figures 5 and 7 provide more robustness checks using different model specifications.

### Results by different air pollutants

We also provide the results using the specific air pollutants (PM_2.5_, PM_10_, SO_2_, NO_2_, CO, O_3_). The results show similar patterns for most of the pollutants: higher pollution levels increase the growth rate of COVID-19 with 2-13 days of delays (Supplementary Figure 8A). For example, a 10% increase in PM_2.5_ (A1), SO_2_ (A3), and CO (A5) statistically significantly leads to a 1.4 ppt, 1.1 ppt, and 3.0 ppt rise in the virus growth rate. The only exception is ozone, for which we find a higher concentration of O_3_ (A6) slightly decreases the disease transmission, even though the relationship is not statistically significant. This might be because (1) the concentration of ozone is often negatively correlated with other pollutants^41^, and (2) ambient ozone can inactivate the virus by disrupting the virus structure^42^. Moreover, across most pollutants, OLS estimates (Supplementary Figure 8B) are substantially smaller than the IV estimates, consistent with our baseline findings.

### Back-of-the-envelope calculations for the “blue sky” policy scenarios

We estimate the excess COVID-19 cases attributable to poor air quality. In China, when daily AQI is below100, it is regarded as “good” or “moderate” air quality (also called a “blue sky” day). During our study period, 18.6% of the city-by-day observations do not meet this standard, and most of the “above-standard” (worse air quality) readings are obtained from northern Chinese cities. Here, we ask what would happen if we were able to bring the air quality index in all Chinese cities to below 100, holding other things constant.

Our estimates imply that the daily virus growth rate would have been slowed by 2.11% on average if all the cities had met the AQI=100 standard (Figure 5A). As a consequence, the number of active infections would have been reduced substantially. For instance, the number of active infections would have dropped from 21,855 to 16,714 (23.5%) on 14 February, when we recorded the highest active infection number in China (Figure 5B). Applying our estimates to the observed removal rate, we expect that the cumulative confirmed cases would have been reduced by 25.7% (30,376 to 22,578) during our study period (Figure 5C).

**Figure 5.**
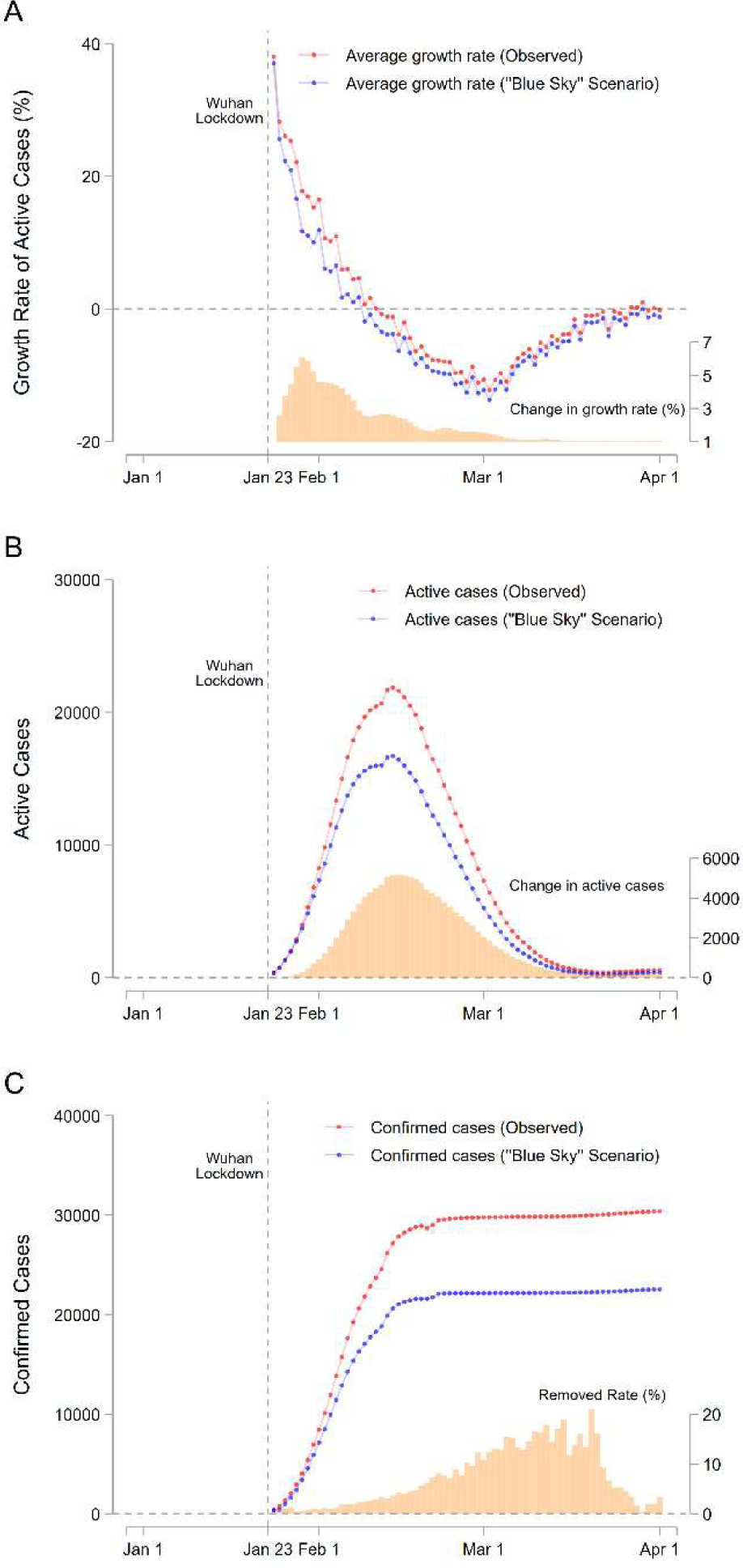
Simulated impacts of air pollution on the daily growth rate, active infections, and confirmed cases of COVID-19. **A**. represents the observed daily growth rate of COVID-19 and the hypothetical rate in the “blue sky” scenario. In the blue sky scenario, an AQI exceeding 100 is replaced with 100. The orange bar represents the change in the growth rate (right axis). **B** shows the active cases in the two scenarios and the orange bar shows the difference. **C**. predicts the estimated cumulative confirmed cases over time. To compute these numbers, we use the observed removal rate, which is shown as the orange bar graphs. See Methods for details.

The simulation shows that air pollution reshaped the exponential growth of the virus. We observe that the difference in the confirmed cases is small in the initial outbreak, but gradually becomes larger as the outbreak progresses. If we do not model the virus transmission process using the SIRD model, we will not be able to capture the dynamic, cumulative, and non-linear impacts of air pollution on COVID-19 cases.

## Discussion

Accurately estimating the effect of air pollution on COVID-19 transmission requires researchers to account for the exponentiality in virus growth and to introduce exogenous shocks to local air quality. Based on the SIRD model, we show that a non-model-based framework could produce biased estimates. This finding suggests that many studies linking air pollution to COVID-19 could be problematic^17-23^. The same concern also applies to studies that try to assess the impacts of virus containment policies^43-45^ and weather conditions^46,47^ on COVID-19 spread. We also documented that associational analysis based on simple OLS regression models will understate the true impact of air pollution on COVID-19 transmission^17,19,21-23^, as is consistent with the previous environmental health literature^24-31^.

Our analyses find that a 10-point (14.3%) increase in AQI leads to a 2.80 percentage point increase in the daily growth rate over 2 to 13 days after the pollution exposure. The effect is statistically significant and economically meaningful. For example, holding all else constant, if we were able to bring all the air quality levels to the country’s standard (AQI = 100, mean AQI would have been reduced by 13.2%), a back-of-envelope calculation reveals that the cumulative cases of COVID-19 would have been reduced by 25.7%

Knowing that air pollution can increase the transmission of COVID-19 is particularly relevant to developing countries that rely heavily on manufacturing and coal (such as India, Indonesia, and Pakistan). These countries face significant challenges in controlling COVID-19, in part because of the faster transmission rate caused by high levels of air pollution. Policymakers should thus consider adopting more stringent pollution control policies in their war to combat COVID-19.

Our study has two caveats. First, we use confirmed active infections to create our outcome variables. As is common in any study of infectious disease^39^, the number of confirmed cases could be much lower than the actual cases, and confirmed cases might not reflect the real epidemic outbreak. This concern is partially alleviated by our use of the growth rate as the outcome variable because our results will hold as long as under-reporting is constant (for example, if 50% of infected cases are always confirmed) within a city. In addition, our regressions include date fixed effects that can control for nationwide events specific to each date, such as national testing policies or revision of the disease classification. While controlling the variation in testing capacity can partially mitigate the concern about under-reporting, such data are not available at the city-by-day level.

Second, we do not have sufficient statistical power to investigate whether air pollution affects COVID-19 deaths. This is because there have been only a few COVID-19 deaths in most Chinese cities. Outside Wuhan, more than 90% of Chinese cities have recorded only 0 or 1 death. While more than 3,000 people died from COVID-19 in Wuhan city, the data were not accurate at the early stage of the outbreak in the city^39^. Therefore, we refrain from discussing the relationship between air pollution and the COVID-19 death rate. Nevertheless, we investigate this issue in Supplementary Figure 9 and Supplementary Note 4 and do not find any statistically significant relationship. Future research is warranted on this issue if richer data from other countries become available.

## Methods

### Materials

#### COVID-19 data

The COVID-19 data are retrieved from the China National Health Commission (CNHC)^48^. The data comprise newly infected, recovered, and death cases from 20 January to 1 April in 2020 in 330 prefectural cities in China. These periods are overlapped with the disease outbreak in China. This includes 49,982 cases and 2,553 deaths in Wuhan and 30,441 cases and 727 deaths in other cities. More than 95% of confirmed cases recovered (tested as negative) during our study period. In our baseline analyses, we exclude Wuhan city due to concerns about the city’s data quality. Wuhan was the epicenter of COVID-19 in China. During the first few weeks after the COVID-19 outbreak, the city faced severe medical resource shortages, and many patients could not get immediate diagnosis and treatment. In fact, a retrospective survey conducted by the Chinese government shows that many COVID-19 deaths in Wuhan may have been misclassified. On 17 April, the Chinese government added another 1,290 COVID-19 deaths in Wuhan (without reporting the timing of the fatalities). COVID-19 data outside Wuhan do not suffer from similar problems because there were far fewer COVID-19 cases in these cities, and COVID-19 tests became widely available soon after scientists learned about the situation in Wuhan.

#### Air quality data

The air quality data are obtained from 1,605 air quality monitoring stations covering all of the prefectural cities in China. These data are available from the Ministry of Ecology and Environment^49^. The AQI is a comprehensive measure of air pollution: the index is constructed using PM_2.5_, PM_10_, SO_2_, CO, O_3_, and NO_2_ concentrations, with a lower AQI meaning better air quality. In China, the AQI is determined by the maximum concentration of different air pollutants. We describe the relationship between the AQI and each pollutant in Supplementary Figure 1 and Supplementary Table 1. To create the city-level air quality data, we first calculated the distance from a city’s population center to all monitoring stations within the corresponding city. We then aggregated station-level air pollution data to city-level data using inverse distance weights. For this process, stations closer to the population center are given higher weights so that city-level air pollution data can better represent the population of each city. The weights are inversely proportional to square distance.

#### Thermal inversion data

The thermal inversion data are obtained from the MERRA-2^50^. The data include the temperature in 42 atmospheric layers (110m to 36,000m). We use the difference in temperature between the first layer (110m) and the second layer (320m) because this is expected to be associated with the air pollution at ground level. The raw data include the information for each 50*60 km grid. We aggregate the grid-level data to the city level using the same methodology as the air pollution data.

#### Weather data

Weather data include temperature, precipitation, and snow depth. These data were obtained from the Global Historical Climatology Network (GHCN) from the U.S. National Oceanic and Atmospheric Administration (NOAA)^51^. We collapse these data to the city by day level using the same method as the air quality data.

## Methods

### SIRD Model

Our empirical analyses are based on the Susceptible-Infectious-Recovered-Deceased Model (SIRD Model), in that the individuals are classified into either Susceptible (*S*_*t*_), Infected (*I*_*t*_), Recovered (*R*_*t*_), or Deceased (*D*_*t*_). The number of each compartment evolves as follows:

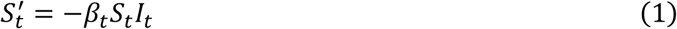

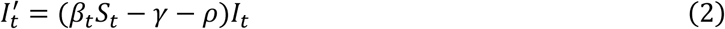

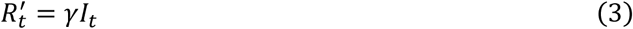

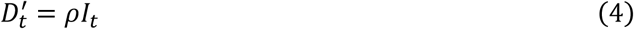

where *β*_*t*_ is transmission rate, *γ* is the recovered rate, and *ρ* is the death rate. Our empirics aim to recover how air pollution affects the transmission rate (*β*_*t*_), which is a widely used parameter to measure the spread of the epidemic because it deterministically affects disease development. Here, the reproduction number (*R*_0*t*_) is proportional to the transmission rate (*R*_0*t*_ = *β*_*t*_/(*γ* + *ρ*)). Using equation (2), the active epidemic growth can be modeled as:

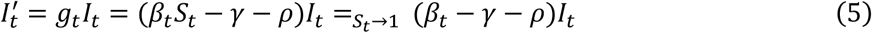

In our analyses, we assume the proportion of the susceptibles is close to 1 (*S*_*t*_ → 1), i.e., almost the entire population can be thought of as susceptible.

The solution of equation (5) can be described by the following exponential function:

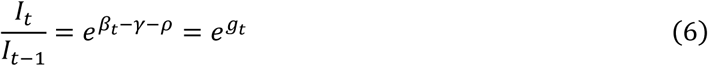

where *I*_*t*_ is active infections at time t, and the infection growth rate (*g*_*t*_) is proportional to the transmission rate (*β*_*t*_). Taking the natural logarithms of this equation, we get

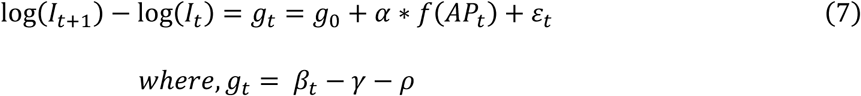

We expect ambient air pollution to alter the COVID-19 growth rate (*g*_*t*_), through changes to the virus transmission rate (*β*_*t*_). Thus, we model the growth rate as a function of air pollution exposure *f*(*AP*_*t*_) and its average treatment effect (*α*), in addition to the baseline growth rate (*g*_0_), which measures the growth rate without any exposure to air pollution, and a mean-zero error term (*ε*_*it*_).

We use the growth rate as the outcome variable because it helps us understand how pollution changes the transmission rate. In contrast, the use of new confirmed cases (or deaths) as an outcome variable, as commonly employed by previous studies^17-23^, could produce estimates that are difficult to interpret. The number of new confirmed cases (*C*_*t*_ − *C*_*t* −1_) could be written as

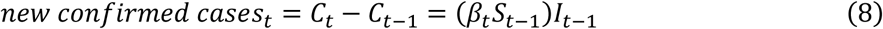

Taking the natural logarithms of this equation, we get

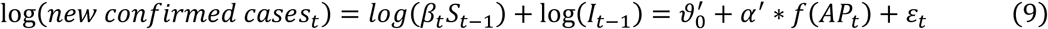

As represented, the number of new confirmed cases depends on both virus transmission rate (*β*_*t*_*S*_*t* −1_) and the current number of active infections (*I*_*t* −1_). Therefore, if we model the new cases as a function of air pollution exposure, the average impact of air pollution (*α*′reflects not only the effect on virus transmission but on the current level of disease spread. Because air pollution could be associated with factors affecting the ongoing disease outbreak, such as the timing of the virus arrival, government interventions, and local economic activities, there could exist spurious correlations between air pollution and these factors.

### Estimation with Flexible Distributed Lag Model

To estimate equation (7), we start by fitting the simple multi-variable regression using Ordinary Least Squares (OLS). The effect of air pollution can appear with lags because the cases will be confirmed after a period of incubation, testing, and reporting. Therefore, we use a Flexible Distributed Lag (FDL) model to allow a dynamic time course of the relationship between air pollution exposure and virus transmission. This can be described as:

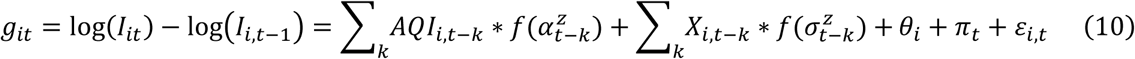

where *g*_*it*_ is the daily growth rate of active infections in city *i* at day *t. AQI*_*i,t*−*k*_ denotes Air Quality Index (AQI) in city *i* at day *t* − *k*, and *X*_*i,t*−*k*_ is a set of control variables in city *i* at day *t* − *k*. We include temperature, precipitation, and snow depth as control variables because these climatic conditions could affect virus transmission^7^. We include the AQI and these control variables during the period between *k* = 0, and *k* = 21 (previous 3 weeks). Existing studies suggest that the virus incubation period is often around 5-6 days, with 2 days being the lower bound^34-36^. Therefore, we are particularly interested in the effect starting from *k* = 2 to *k* = 13. Indeed, in our estimation, the impact during this window is the most salient, and including more periods in the regressions does not change the magnitude of the effect but enlarges its standard errors.

*α*_*t*−*k*_ represents the effect of air pollution on the daily virus growth rate in period *t-k*. To investigate the dynamic impact of air pollution, we adopt the FDL model, which approximates the set of coefficients *α*_*t*−*k*_ as a cubic B-spline function with z segments, as denoted by 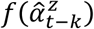. Because daily variation in air pollution generally is highly correlated, this smoothing process helps us reduce artificial oscillations from our parameter estimates and interpret the coefficients easily; without this process, we might observe some random spikes and descents in the estimated parameters, which could not be rationalized^38^. In our primary model, we adopt z = 3 segments. For the robustness check (Supplementary Figures 4 and 7), we also choose z = 2 and z = 4.

*θ*_*i*_ and *π*_*t*_ denote city and time fixed effects, which are a set of city-specific and time-specific dummy variables. The inclusion of these sets of fixed effects helps isolate variation in air pollution exposure from time-invariant, time-trending, or seasonal confounders, which could be correlated with virus transmission. Specifically, city fixed effects (*θ*_*i*_) account for time-invariant confounders specific to each city (e.g., the city’s income level, natural endowments, and short-term industrial and economic structure), while time fixed effects (*π*_*t*_) account for shocks that are common to all cities on a given day (e.g., national virus containment policies, macroeconomic conditions, and the national air pollution time trend). Conditional on these fixed effects and the time-varying control variables, we assess how air pollution affects virus growth rate. If air pollution accelerates virus transmission, *α*_*t*−*k*_ will be positive.

Note that, as is common in any infectious disease literature, the confirmed active cases could be substantially lower than the true cases, and the use of the confirmed cases might not be precise^39^. In our study, this concern can be partially alleviated for at least three reasons. First, we use the growth rate of confirmed active infections as our outcome variable, so our findings will not be affected as long as the under-reporting is constant (for example, 50% of infections are always confirmed) within a city. Second, our regression includes date fixed effects, and this can absorb the national-level testing policies or revision of the disease classification. For example, the national government changed the disease classification on 18 February, but this variation can be absorbed by including date fixed effects. Finally, we exclude Wuhan, in which misreporting was more frequent^39^.

### Two-Stage Least Squares Strategy

Isolating the effect of air pollution from other confounding factors that could also affect the virus spread is a central empirical challenge in estimating the causal relationship between air pollution and COVID-19 transmission. In the literature, it is well documented that omitted variables and measurement errors could under-estimate the true pollution effects on health (also see Supplementary Figure 6)^24-31^. In addition, there are at least two additional potential sources of endogeneity when investigating the effects on infectious diseases.

First, the variation in air pollution could be correlated with individuals’ averting behaviors, which could also affect virus transmission. When people are aware of severe air pollution, they might wear masks, purchase air purifiers, or reduce outdoor activities^52,53^, which could reduce virus transmission. If this is the case, the OLS estimators might understate the true impact of air pollution. Second, economic activities (e.g., opening industries and schools), human mobility (e.g., use of transportation), and health interventions (e.g., social distancing and business closures) can affect the disease spread, and these could also be correlated with air quality^,33^. This implies that even if we observe a positive association between air pollution and the epidemic’s growth (with potential delays), the association might not necessarily reflect the pollution-disease relationship.

Our solution to address these empirical threats is to adopt the Instrumental Variable (IV) approach. We use thermal inversion to instrument air pollution^25,34^. Thermal inversion is a natural phenomenon in which a layer of cooler air is overlaid by a layer of warmer air in the atmosphere. When a thermal inversion occurs, the air pollutants emitted from the ground surface will be trapped, which raises the air pollution concentration. We use a 2SLS procedure to estimate the IV model. The first stage in 2SLS can be described as

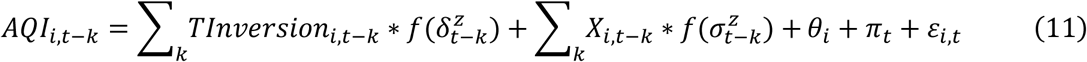

where *TInversion*_*i,t*−*k*_, denotes the difference in temperature between surface ground and upper layer in city *i* in time *t-k*, with the larger number representing the warmer temperature in the upper layer. (Therefore, we expect 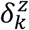 to be positive.) Other variables are analogous to equation (10).

In the second stage, we regress the daily virus growth rate on this predicted pollution concentration rather than on observed pollution concentration using the following equation:

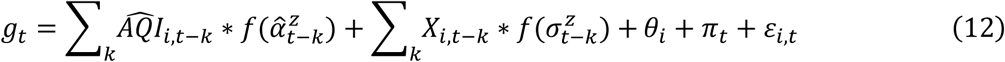

where 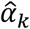 measures the plausibly causal impact of air quality on virus transmission. In other words, the thermal inversion-driven air pollution has to be uncorrelated with the error term *ε*_*i,t*_, conditional on the set of fixed effects and time-varying control variables, which is not directly testable. Here, the inclusion of these control variables can help isolate the variation in air pollution from other confounders at the cost of reducing much of the variation in pollution exposure. We cluster the standard errors at the city level.

### Placebo Test

To examine the relationship between thermal inversion and air pollution, we regress daily air pollution levels on the occurrence of thermal inversions, conditional on the set of control variables and fixed effects used in equation (10) - (12). We further conduct a “placebo” test to rule out the possibility that the strong inversion-pollution relationship is driven by the local trends/seasonality, the spatial distribution of frequency of the thermal inversion, or other factors. Specifically, we randomly shuffle the observed thermal inversions within the same location or within the same day by 1000 times and re-estimate the relationship between pollution and placebo inversions. We plot the distribution of the estimated coefficients and find that their average effects are close to zero.

### Back-of-the-envelope calculation

We estimate the excess COVID-19 cases attributable to poor air quality. Developing countries, including China, have suffered greatly from poor environmental quality as the cost of rapid economic growth^54^. In China, AQI lower than 100 is regarded as “moderate” or “good” air quality (See Supplementary Table 1) and is recognized as the “blue sky”. However, during our study period, 18.6% of the samples (city-by-day) did not meet this standard, and 30% of the cities explains 72.9% of the share with poor air quality. To estimate the excess COVID-19 cases, we first estimate their daily growth rate in two scenarios: observed air quality and the blue sky scenario (AQI is always lower than 100) by calculating the following equations.

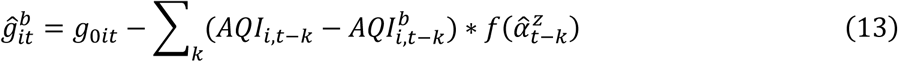

where *g*_0*it*_ denotes the observed daily growth rate in city *i* at time *t*, and 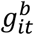 denotes the growth rate if the AQI were always lower than 100. *AQI*_*i,t*−*k*_ is observed AQI and 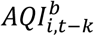 is the hypothetical AQI, in that AQI higher than 100 is replaced with 100. 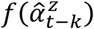 represents the estimated coefficients from our main estimates (equation (12)). As discussed in the main analyses, we only consider impacts between k = 2 and k = 13.

Using the predicted growth rate, the active infections in the blue sky scenario would have been:

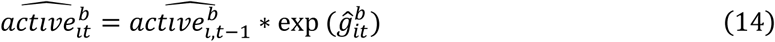

The predicted number of active infections in city *i* at time *t* builds on that number in the previous period. To predict these numbers, we use the initial active cases in each city when it exceeds 10. We then use the observed removal rate in equations (3) and (4) to estimate the cumulative confirmed cases.

For the removal rate, we use the following equation.

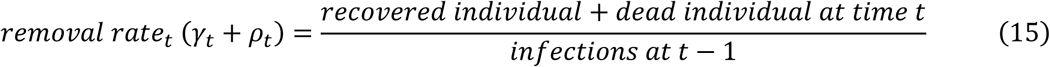

For this projection, we allow the removal rate to vary over time because it should vary depending on the stage of the outbreak. Note that the individuals are classified as recovered when they are tested as negative in our dataset. Therefore, its definition is slightly different from that in the classical SIRD model (See Supplementary Note 3).

## Data Availability

The data will be made available upon the publication of the paper.

## Acknowledgment

We would like to thank Wenbo Gong, Yatang Lin, and Deyu Rao for their valuable comments and suggestions. TT also thanks the Bai Xian Asia Institute for the scholarship support as a Bai Xian Scholar.

## Author Contributions

All authors (GH, PY, and TT) contributed equally to the paper. GH, PY, and TT conceptualized the study and carried out initial planning. PY retrieved and constructed the data set. PY and TT carried out the statistical analysis, which was refined by GH for the final version. TT prepared the first draft of the report, which was revised by PY and GH. All authors reviewed and contributed to a final draft and approved the final version for publication.

## Data and Code Availability Statement

All data and code necessary for replication will be available at a public repository upon publication.

## Supplementary Materials

### Supplementary Notes

#### Supplementary Note 1: Mechanisms of air pollution and COVID-19 transmission

In this note, we elaborate how air pollution could affect COVID-19 virus transmission. First, short-term exposure to severe air pollution can weaken the immune system’s response to the virus and thereby increase the chance of infection^55,56^. In physiological studies, inhaled particulates and other air pollutants can interact with immune cells within the airways. In particular, it is found that pollutants can trigger cellular signaling pathways, leading to multicellular immune responses and perturbation, and eventually cause disease or fail to prevent disease^57,58^. In epidemiological studies, air pollutants such as PMs, NO_2_, SO_2_, and O_3_ are also found to be associated with inflammatory and immune responses^59,60^.

Second, aerosols in air pollutants may maintain virus activity and facilitate virus transmission. Aerosols are suspensions of solid or liquid particles in the air. These particles are small and have a low settling velocity; therefore, they can remain airborne for prolonged periods. For example, coughing and sneezing can generate a substantial quantity of particles. Because of evaporation, a large number of these particles shrink and then behave as aerosols. Aerosols due to coughing, sneezing and breathing can result in virus transmission, in which the virus from an infected person can be carried over a fairly long distance, compared to transmission through large droplets and direct contact. Both laboratory and epidemiological studies have shown that aerosol transmission can be an important mode of transmissions of influenza, chickenpox, measles, tuberculosis, smallpox, H5N1, MERS, Ebola^61-66^, and SARS-CoV-2 (i.e., COVID-19) ^11-13^.

#### Supplementary Note 2: COVID-19 Lockdown Data

We collected local governments’ lockdown information city by city from various news media and government announcements (Supplementary Table 3). Most of the cities’ lockdown policies were directly issued by the city-level governments, although a few were promulgated by the provincial governments. To ensure compliance, civil servants and volunteers were assigned to communities, firms, business centers, and traffic checkpoints. Local governments also penalized offenders if the rules were violated. There were some variations in rules and in the degree of the lockdown. For example, in some cities, individuals were not allowed to go out (food and daily necessities were delivered to them), while in other cities, they could go out if they did not have a fever. In this paper, we designated a city as locked down when the following three measures were all enforced: (1) prohibition of unnecessary commercial activities in people’s daily lives, (2) prohibition of any type of gathering by residents, (3) restrictions on private vehicles and public transportation. Following our definition, 95 out of 324 cities were locked down in our study period.

#### Supplementary Note 3: Removal rate

For the back-of-the-envelope calculation, we use the observed removal rate to consistently project the epidemic growth in different policy scenarios. However, in our dataset, the individuals with infections are classified as recovered when they test negative, rather than when they recover from symptoms. Therefore, to estimate the reproduction number from our main results, we adopt the removal rate which is reported by existing studies. They suggest that an average interval is 5.1 days^34^, 5.8 days^35^, or 7.5 days^36^, making the removal rate 13.3%-19.6%. Therefore, we use a removal rate between 13%-20% to compute the reproduction number (*R*_0_ = *β*/(*γ* + *ρ*)).

#### Supplementary Note 4: The Effect of Air Pollution on the COVID-19 Death Rate

Existing studies suggest that air pollution increases deaths from COVID-19, likely because air pollution could impair the physical capability to recover from the infection. This is policy-relevant, but our dataset does not have sufficient statistical power to investigate the relationship. Outside Wuhan, there have been only around 700 deaths from the pandemic, with about 90% of cities having only 0 or 1 death. While more than 3,000 deaths are recorded in Wuhan, the data may not be credible. For example, on 17 April, its official death record was revised, with 1,290 deaths being added but without the timing of the deaths.

Nevertheless, here we examine the relationship between air pollution and death rate by aggregating the daily level data to the weekly level so that we can have a wider variation in the outcome. Our outcome variable is not the growth rate of deaths because it should be proportional to the growth rate of the infections. Instead, we use the death rate, which is defined as the probability of death given infection. Using this outcome variable, we fit the following model, analogous to equation (7),

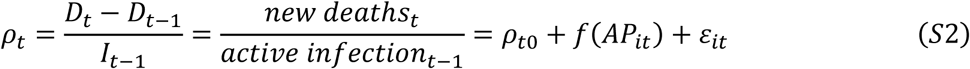

where the death rate is a function of baseline death rate, air pollution, and mean zero error term. We use the same control variables and a set of fixed effects as in equations (9) - (11).

We do not find any suggestive evidence that air pollution increases the death rate of the patients. All coefficients seem to be very small, and the sign is not consistent. As expected, our data may not have sufficient statistical power to investigate the relationship accurately.

**Supplementary Figure 1.**
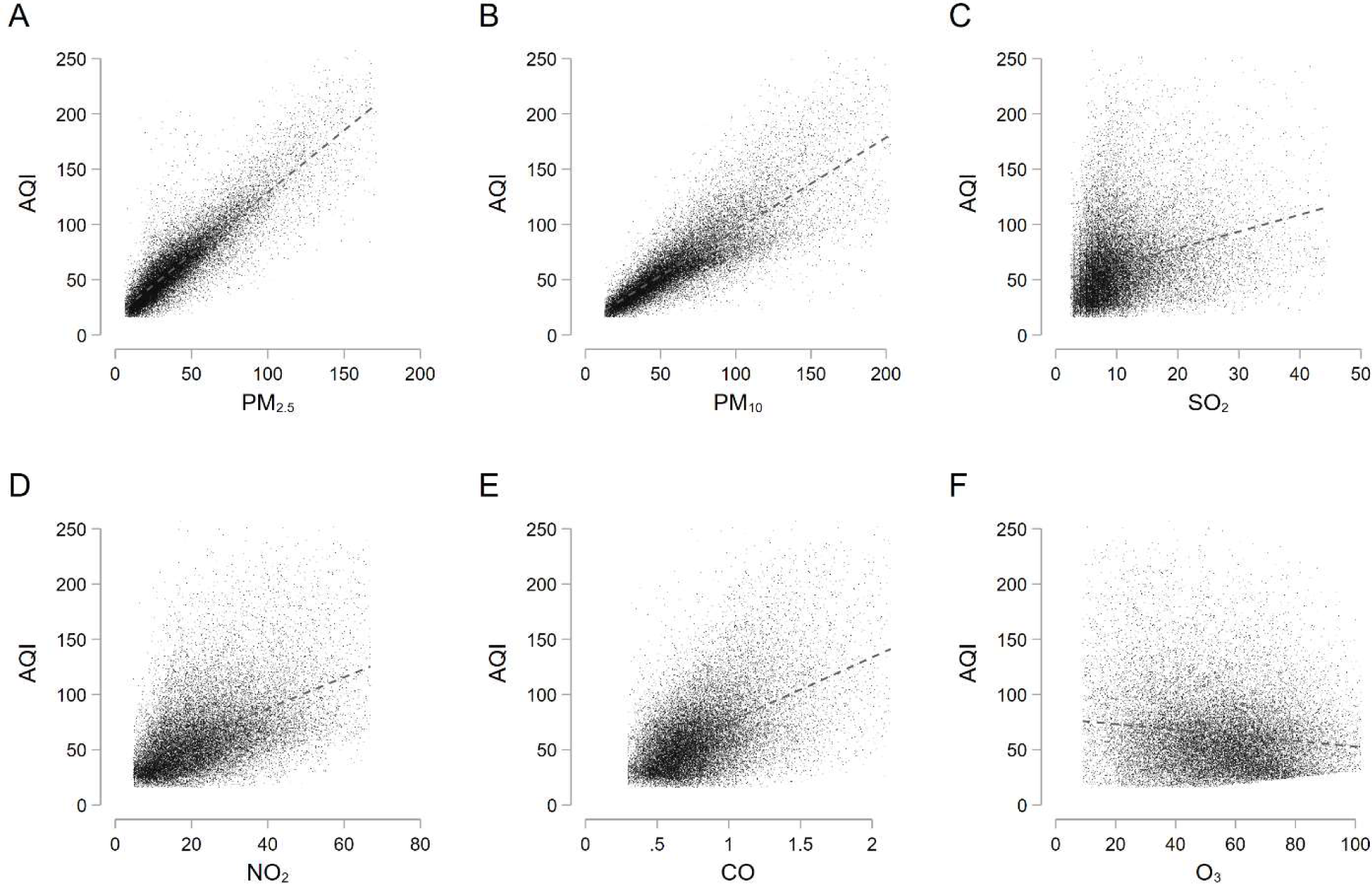
Air Quality Index is correlated with the most important pollutants. A-F represents the relationship between the AQI and each pollutant, including PM_2.5_, PM_10_, SO_2_, NO_2_, CO, and O_3_, which are components of the index. During the wintertime in China, PM_2.5_, PM_10_ domain the primary pollutants of AQI in most cities. There is a strong correlation between the AQI and each pollutant, except for O_3_ (ozone). This might be because ozone is mechanically negatively correlated with some of the primary pollutants. We trim the observations below 1 percentile and above 99 percentile in each pollutant. See Extended Data Table 1 for the definition of the AQI.

**Supplementary Figure 2.**
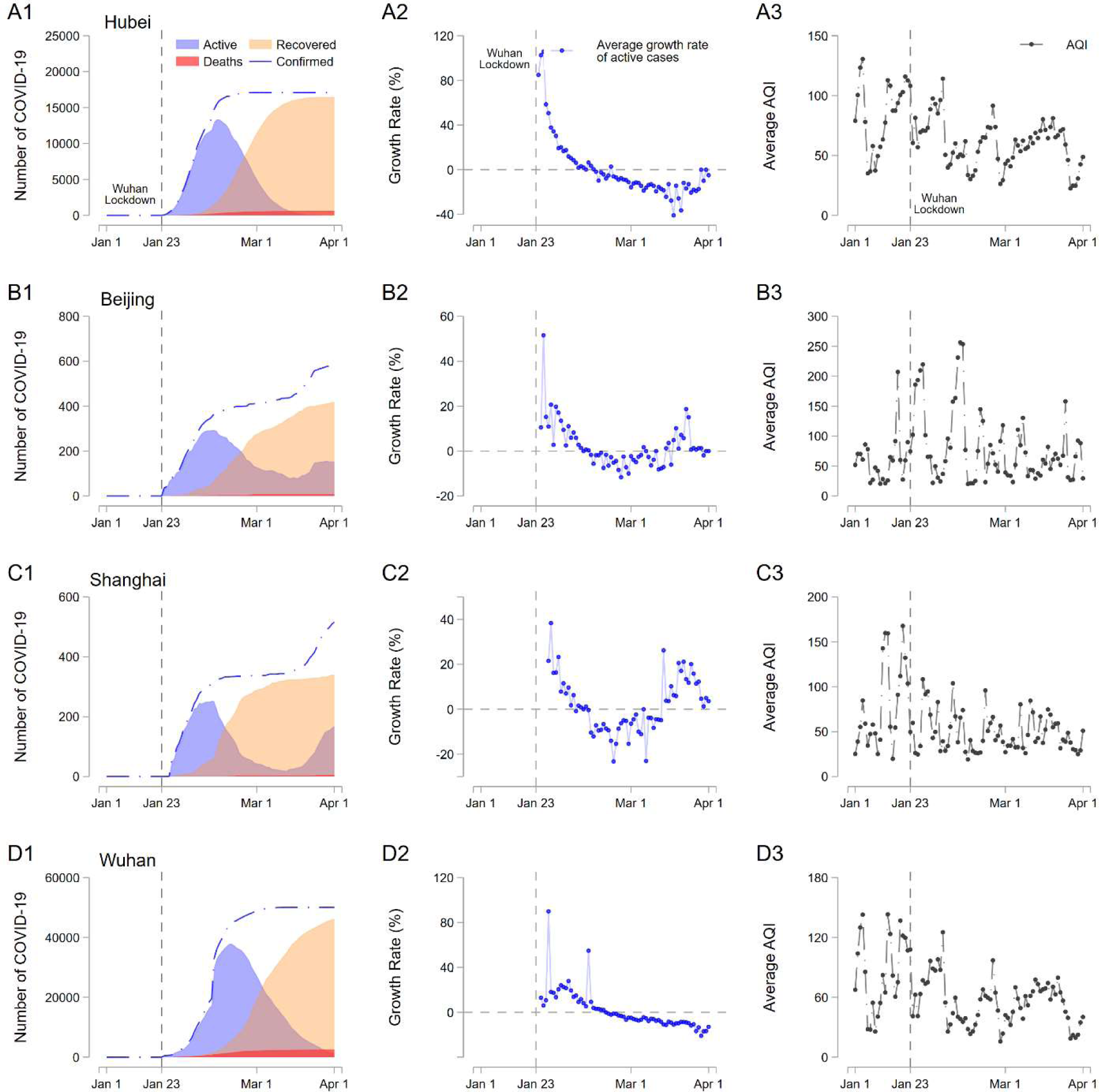
COVID-19, its growth rate, and the Air Quality Index in Hubei Province, Beijing, Shanghai, and Wuhan. These graphs show the COVID-19 outbreak (number of active, recovered, and deceased from COVID-19), the infection growth rate, and AQI in each region.

**Supplementary Figure 3.**
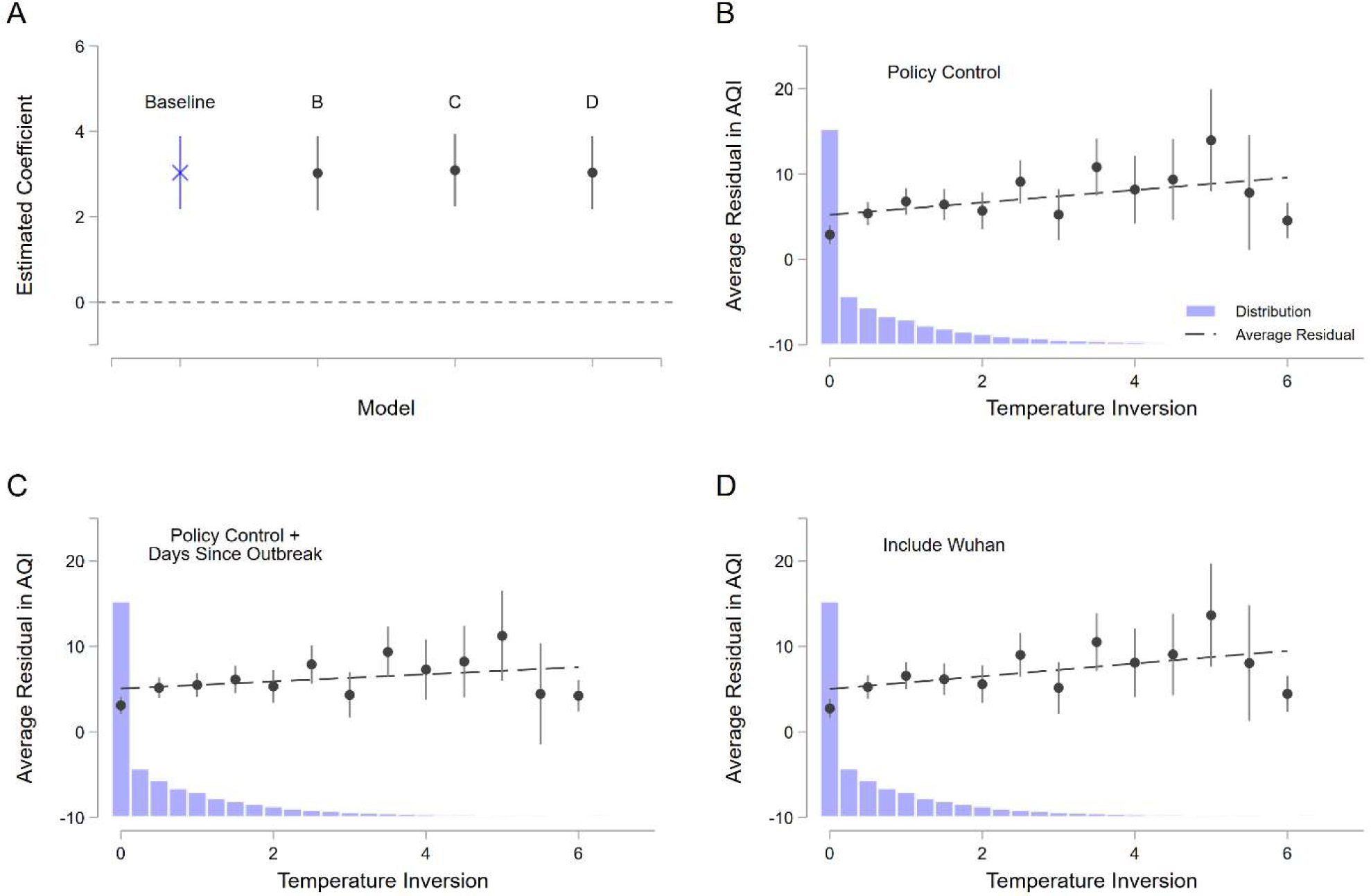
Strong correlation between thermal inversion and air pollution is robust to different specifications. These graphs show the correlation between temperature inversion and residual in the Air Quality Index, after controlling for the weather variables, city fixed effects, and date fixed effects. **A**. shows the coefficients of the contemporaneous relationship for each model, which corresponds to Figure 3A. **B-D**. show the distribution of thermal inversion and the average residual. **B**. includes lockdown status for the control variable, **C**. includes lockdown status and days since the outbreak (first confirmed cases). **D**. includes Wuhan city in the regression.

**Supplementary Figure 4.**
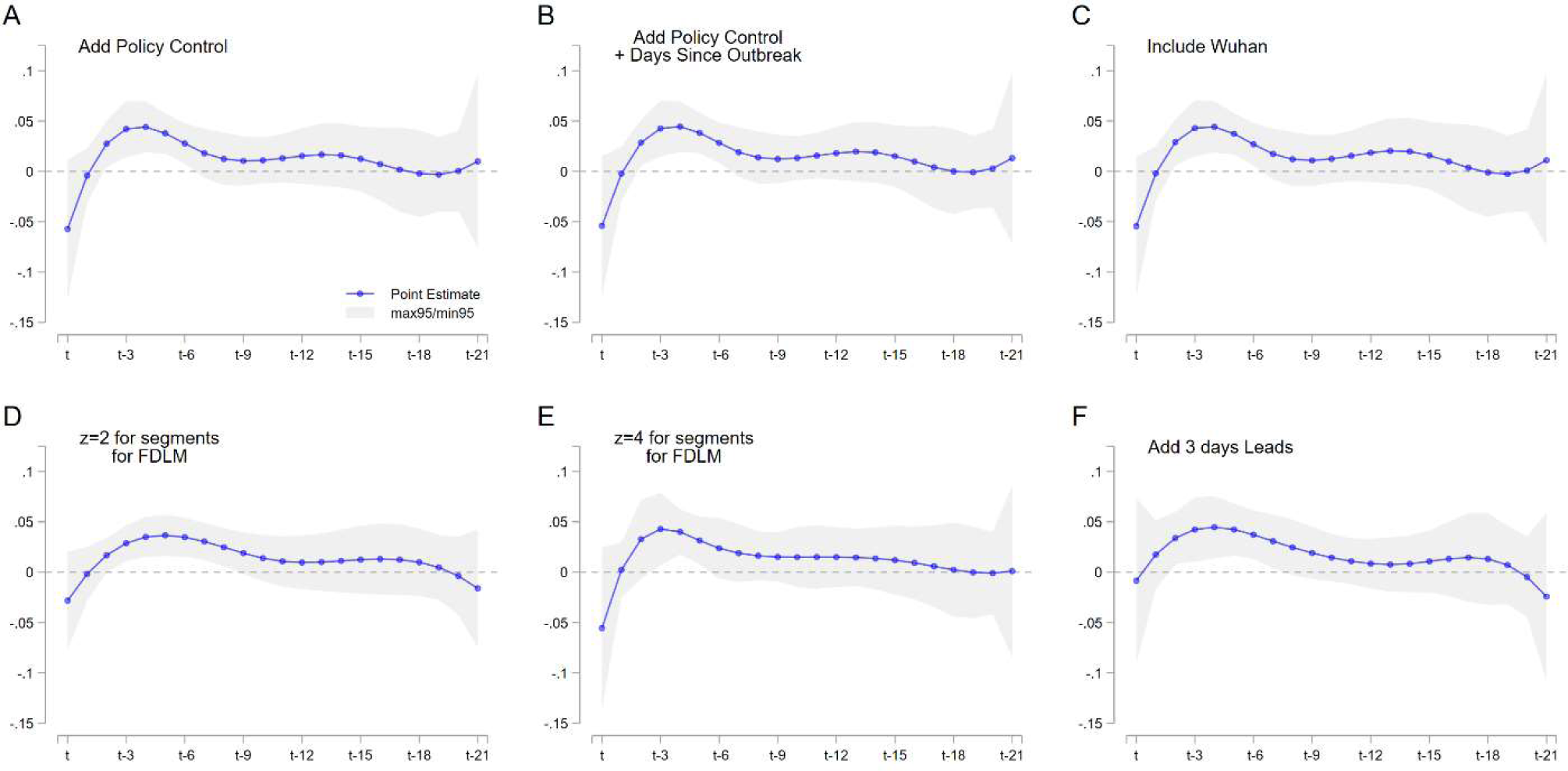
The effects of air pollution on the COVID-19 growth rate using the IV estimates are robust to a number of model specifications. **A**. lockdown status is added as a control variable. **B**. the lockdown status and days since the outbreak (the first case confirmed) are added in the regression. **C**. We include Wuhan. **D and E**. We adopt different segments for the Flexible Distributed Lag Model. **F**. We add three days of future air pollution. The joint coefficient for the three days lead is 0.002, with the standard error at 0.051, suggesting that future air pollution does not affect the disease growth rate. Thermal inversions and the same controls are used for all first-stage regressions. All regressions include weather controls (temperature, precipitation, and snow depth), date fixed effects, and city fixed effects. Standard errors are clustered at the city level.

**Supplementary Figure 5.**
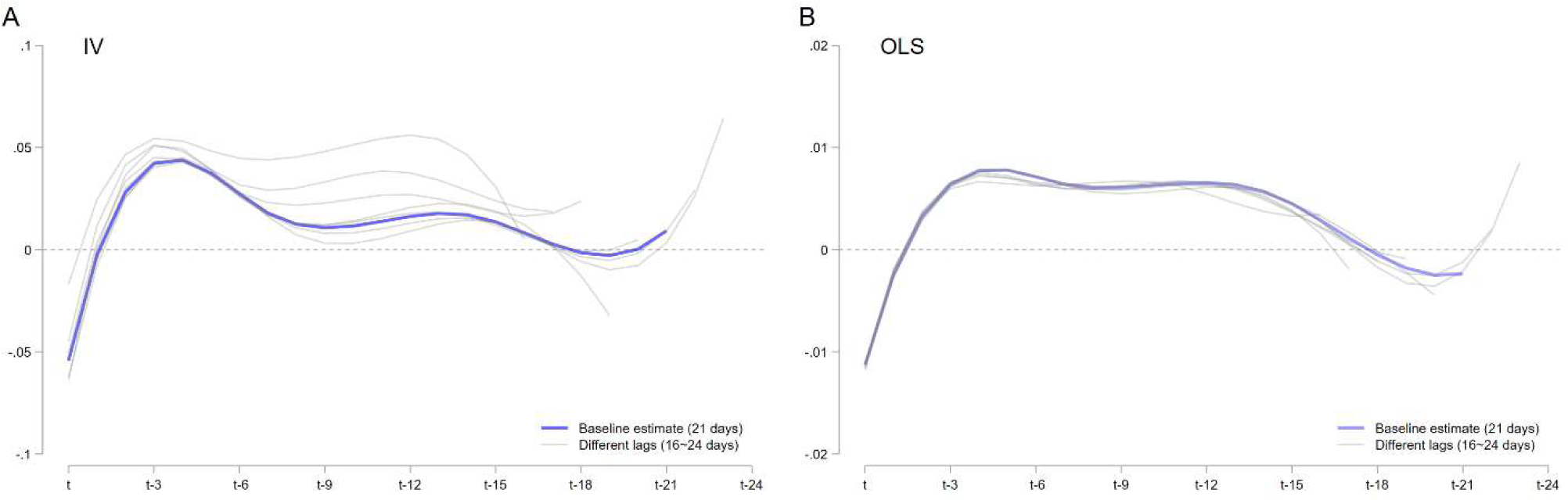
The effects of air pollution on the COVID-19 growth rate using different lags. **A**. represents the results using IV. We use different lengths of lags from 16 days to 24 days, while the main estimation uses 21 days. The blue line shows the baseline estimates, and the gray line shows the results with different lags. **B**. shows the results using OLS estimates.

**Supplementary Figure 6.**
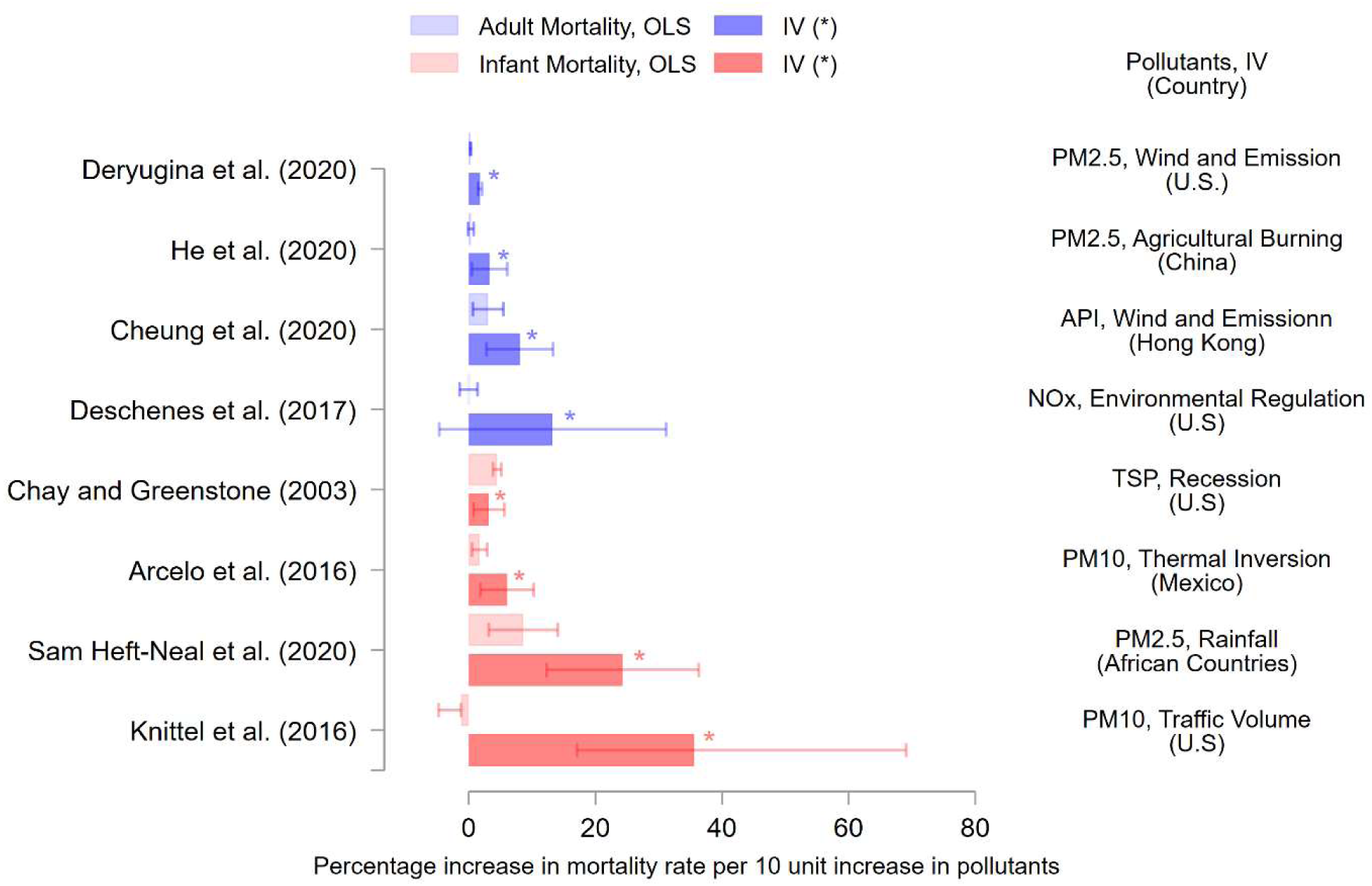
Instrumental variables (IV) estimates are consistently larger than OLS estimates in existing studies linking air pollution and health outcomes. The graph represents the estimates of the effect per 10 unit increase in air pollution on mortality rate (%). Deryugina et al. (2020) use mortality among aged above 65. Except for Chay and Greenstone (2003), existing studies report the OLS estimates are smaller than the IV estimates. Note that different studies focus on different pollutants and use different instrumental variables. Therefore, we do not compare estimates across different studies but across different methods within each study.

**Supplementary Figure 7.**
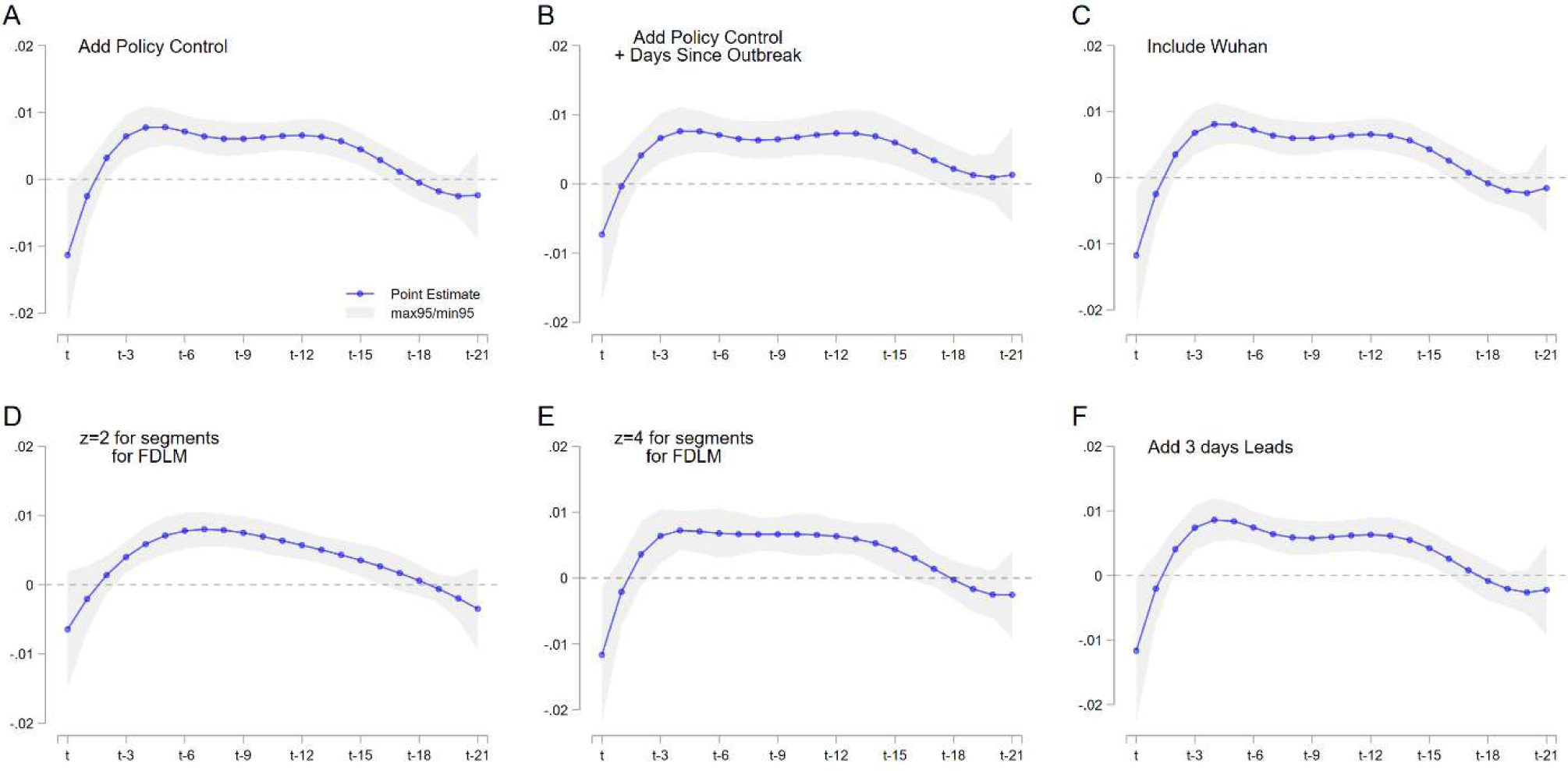
The correlations between air pollution and COVID-19 growth rate using OLS estimates are robust to a number of model specifications. **A**. lockdown status is added as a control variable. **B**. the lockdown status and days since the outbreak (the first case confirmed) are added in the regression. **C**. We include Wuhan. **D and E**. We adopt different segments for the Flexible Distributed Lag Model. **F**. We add three days of future air pollution. The joint coefficient for the three days lead is 0.010, with the standard error at 0.010, suggesting that future air pollution does not affect the disease growth rate. All regressions include weather controls (temperature, precipitation, and snow depth), date fixed effects, and city fixed effects. Standard errors are clustered at the city level.

**Supplementary Figure 8.**
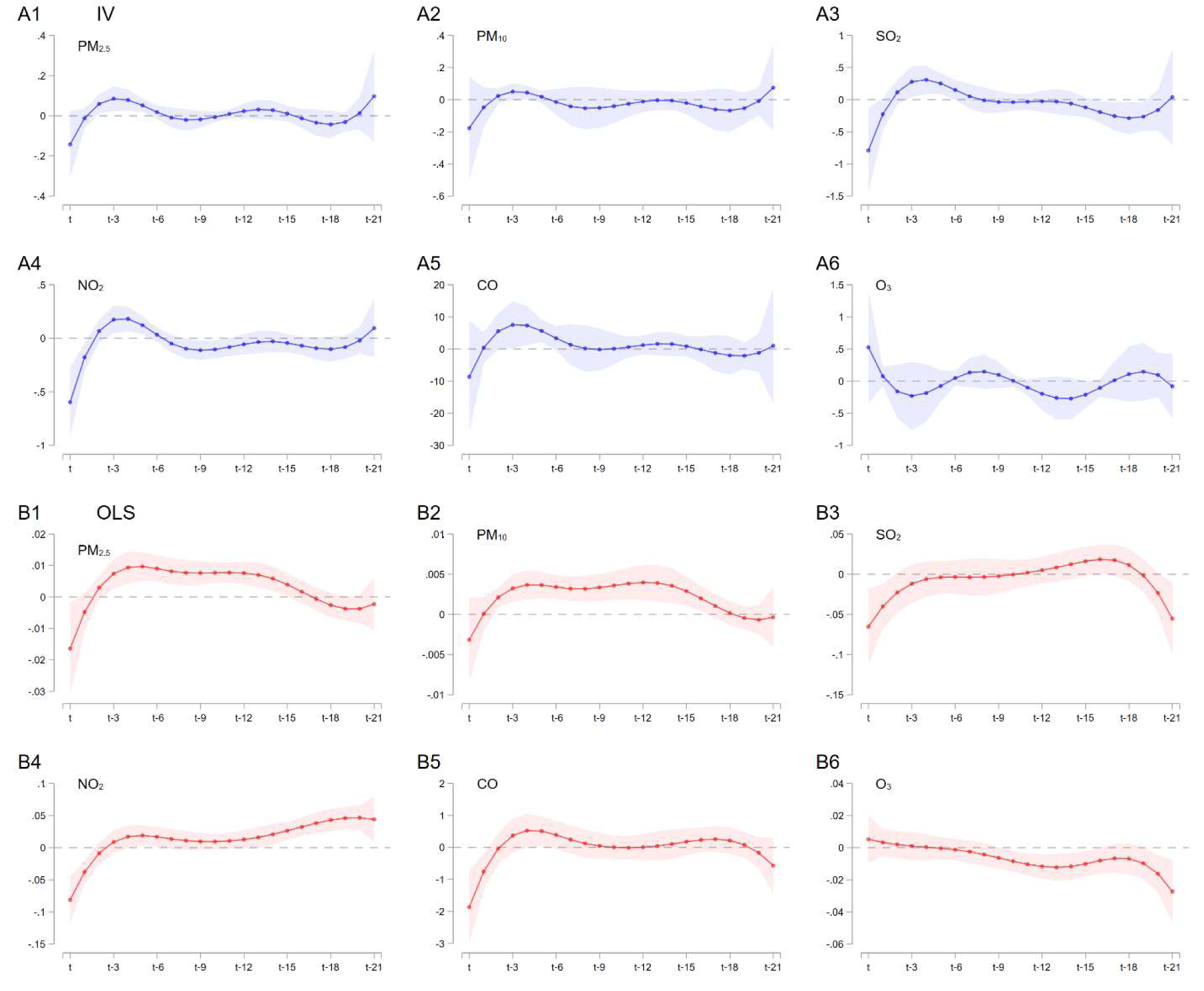
The effects of air pollution on the COVID-19 growth rate using different air pollutants. **A1-A6**. Represents the results using IV. Each graph shows the coefficient of a 1 point increase in each pollutant in each day. Weather controls (temperature, precipitation, and snow depth), date fixed effects, and city fixed effects are included in both the first and second stage regression. The blue line represents the point estimates, while the blue area denotes the 95% confidence interval. **B**. represents the results using OLS. The regression includes the same controls as the IV estimates. The red line represents the point estimates, while the red area denotes the 95% confidence interval. In all regressions, standard errors are clustered at the city level.

**Supplementary Figure 9.**
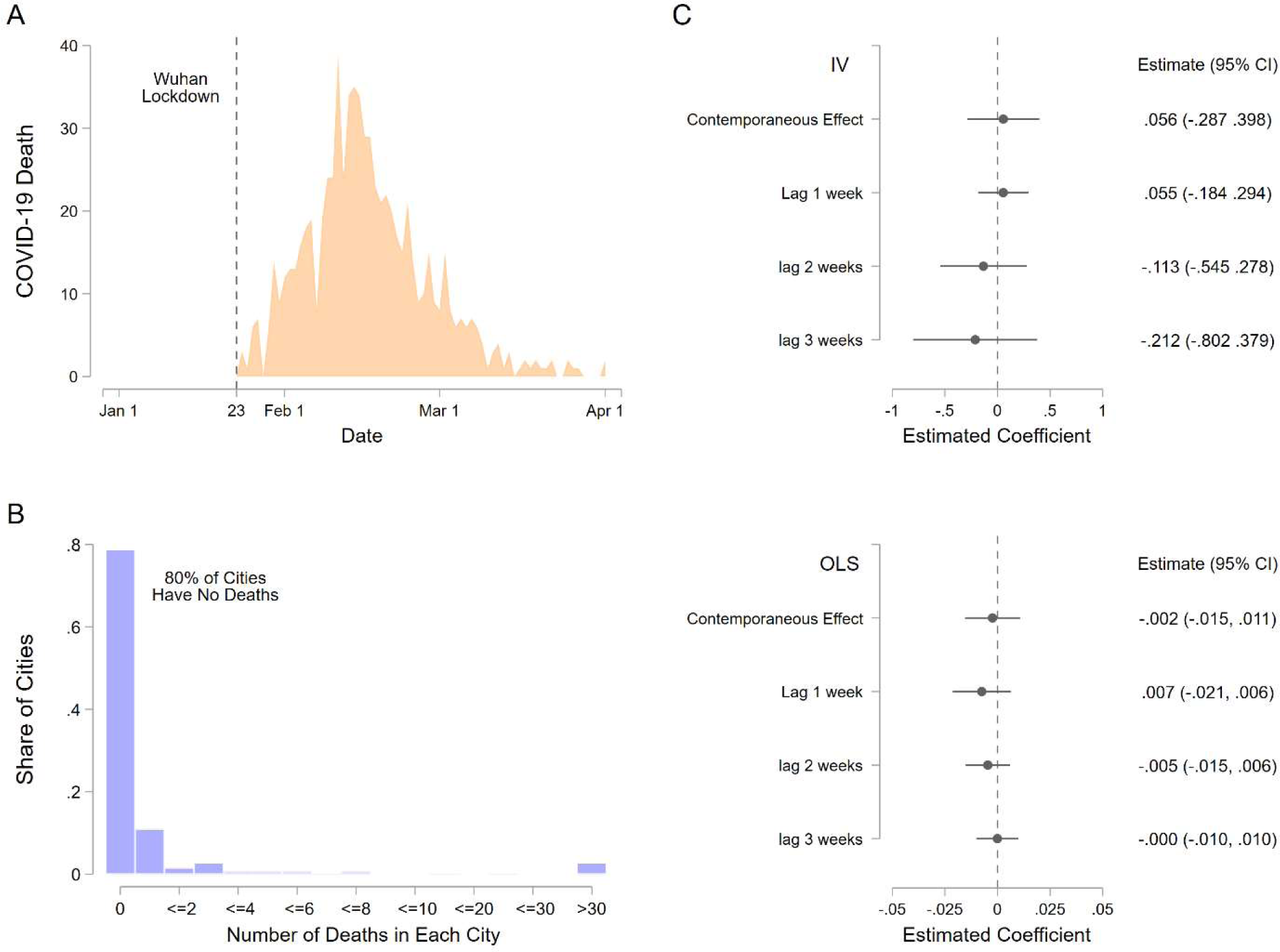
Impacts of air pollution on the COVID-19 death rate. **A**. The trend of the COVID-19 deaths over time outside Wuhan. **B**. Distribution of deaths in each city. **C**. The graph above shows the effect of air pollution on death rate using IV, while the graph below shows the effect using OLS. To keep the variation in death rate, we aggregate data to the week-by-city level. The results do not show a statistically significant relationship between AQI and the death rate.

### Supplementary Tables

**Supplementary Table 1.**
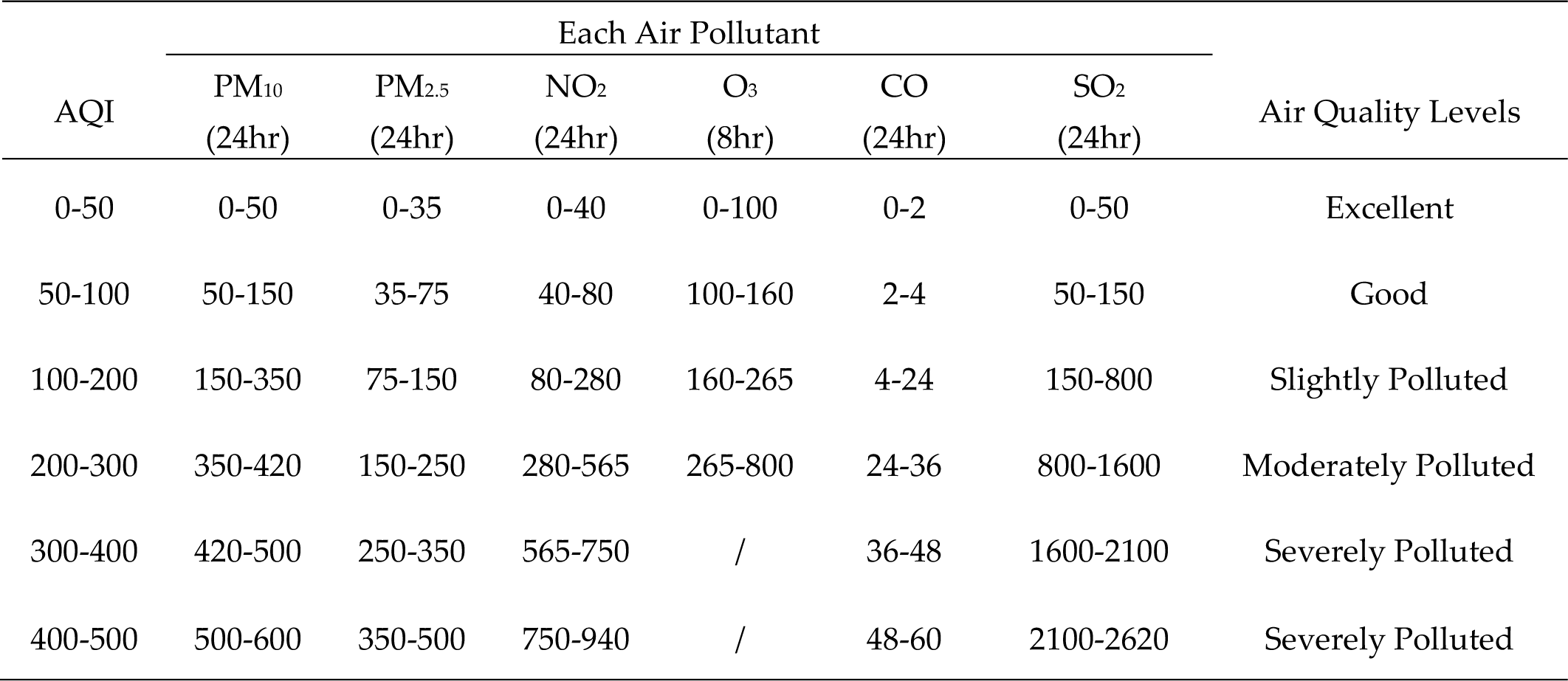
The relationship between the AQI and different air pollutants. This table reports the AQI sub-index levels for each air pollutant. The sub-index with the highest value will then be used as the AQI. For CO, the unit is mg/m^3^, and for other pollutants, the units are µg/m^3^.

**Supplementary Table 2.** Full results of the effect of air quality on the COVID-19 growth rate. The results correspond to Figure 4. The dependent variable is the day-by-city level growth rate of the activated COVID-19 cases. Each estimate indicates the effect of the current and past air pollution (Air Quality Index) on the growth rate of COVID-19. Weather controls include temperature, precipitation, and snow depth. Standard errors are clustered at the city level and shown in the right-side brackets. Significance levels are indicated by *** p<0.01, ** p<0.05, and * p<0.01.

| Dependent Variable:<br>Daily disease growth rate | OLS |  | IV |  |
| --- | --- | --- | --- | --- |
|  | coefficients | s.e. | coefficients | s.e. |
|  | (1) | (2) | (3) | (4) |
| Current Day | -0.011** | [0.005] | -0.054 | [0.035] |
| lag: 1 day | -0.003 | [0.002] | -0.002 | [0.014] |
| lag: 2 days | 0.003** | [0.002] | 0.028** | [0.012] |
| lag: 3 days | 0.006*** | [0.002] | 0.042*** | [0.014] |
| lag: 4 days | 0.008*** | [0.002] | 0.044*** | [0.013] |
| lag: 5 days | 0.008*** | [0.001] | 0.038*** | [0.010] |
| lag: 6 days | 0.007*** | [0.001] | 0.028*** | [0.010] |
| lag: 7 days | 0.006*** | [0.001] | 0.018 | [0.013] |
| lag: 8 days | 0.006*** | [0.001] | 0.012 | [0.013] |
| lag: 9 days | 0.006*** | [0.001] | 0.011 | [0.013] |
| lag: 10 days | 0.006*** | [0.001] | 0.012 | [0.012] |
| lag: 11 days | 0.006*** | [0.001] | 0.014 | [0.012] |
| lag: 12 days | 0.007*** | [0.001] | 0.016 | [0.014] |
| lag: 13 days | 0.006*** | [0.001] | 0.018 | [0.016] |
| lag: 14 days | 0.006*** | [0.001] | 0.017 | [0.016] |
| lag: 15 days | 0.004*** | [0.001] | 0.014 | [0.016] |
| lag: 16 days | 0.003** | [0.001] | 0.008 | [0.019] |
| lag: 17 days | 0.001 | [0.001] | 0.003 | [0.021] |
| lag: 18 days | -0.000 | [0.001] | -0.001 | [0.022] |
| lag: 19 days | -0.002 | [0.001] | -0.003 | [0.019] |
| lag: 20 days | -0.002 | [0.002] | 0.000 | [0.020] |
| lag: 21 days | -0.002 | [0.003] | 0.009 | [0.044] |
| z-Segment, k-order | 3, 3 |  | 3, 3 |  |
| Observations (cities) | 22,701 (329) |  | 22,701 (329) |  |
| Weather Control | Y |  | Y |  |
| Date fixed effects | Y |  | Y |  |
| City fixed effects | Y |  | Y |  |

**Supplementary Table 3.**
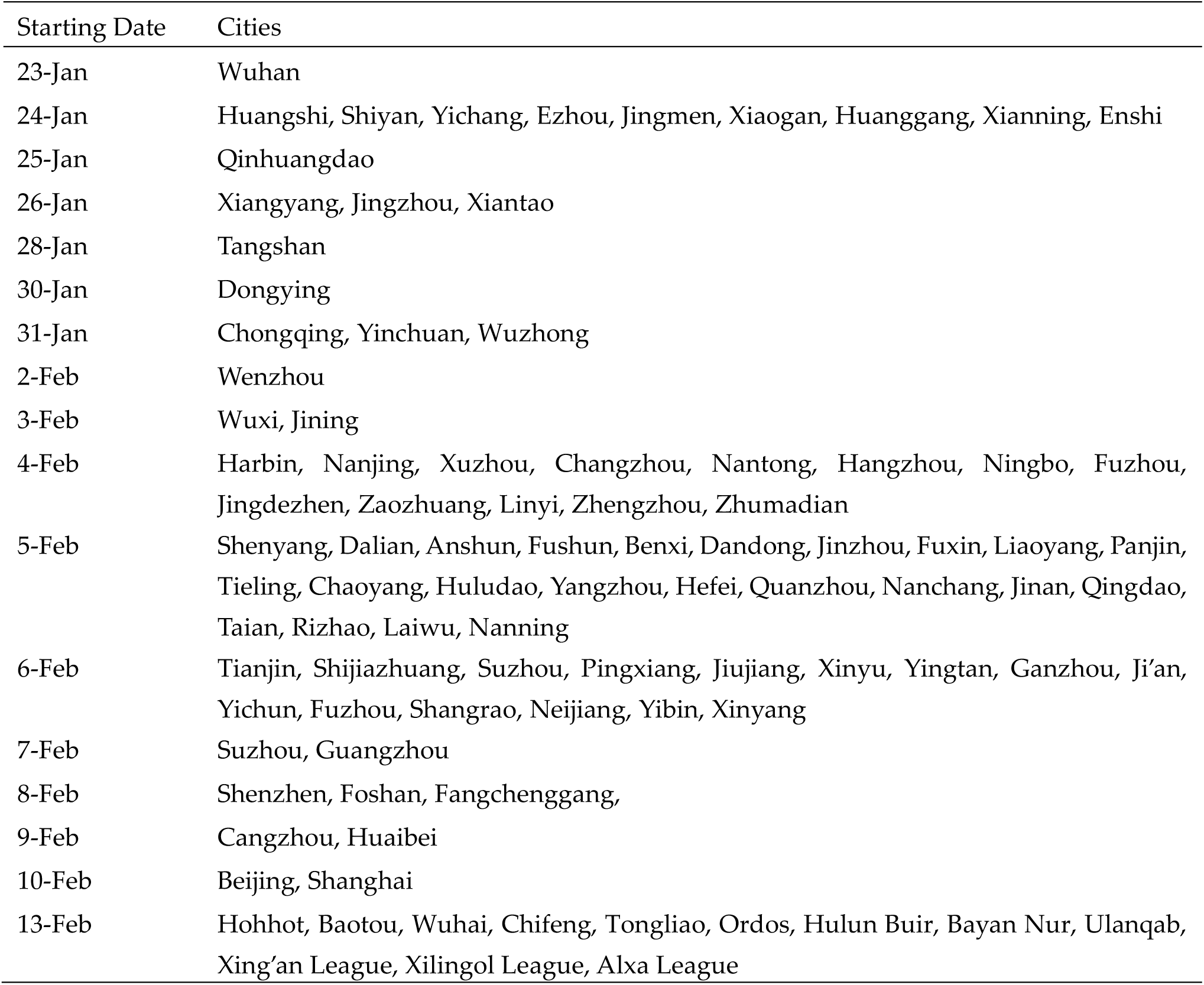
List of locked-down cities. The lockdown information is from local government and various media news in 2020.

| Starting Date | Cities |
| --- | --- |
| 23-Jan | Wuhan |
| 24-Jan | Huangshi, Shiyan, Yichang, Ezhou, Jingmen, Xiaogan, Huanggang, Xianning, Enshi |
| 25-Jan | Qinhuangdao |
| 26-Jan | Xiangyang, Jingzhou, Xiantao |
| 28-Jan | Tangshan |
| 30-Jan | Dongying |
| 31-Jan | Chongqing, Yinchuan, Wuzhong |
| 2-Feb | Wenzhou |
| 3-Feb | Wuxi, Jining |
| 4-Feb | Harbin, Nanjing, Xuzhou, Changzhou, Nantong, Hangzhou, Ningbo, Fuzhou, Jingdezhen, Zaozhuang, Linyi, Zhengzhou, Zhumadian |
| 5-Feb | Shenyang, Dalian, Anshun, Fushun, Benxi, Dandong, Jinzhou, Fuxin, Liaoyang, Panjin, Tieling, Chaoyang, Huludao, Yangzhou, Hefei, Quanzhou, Nanchang, Jinan, Qingdao, Taian, Rizhao, Laiwu, Nanning |
| 6-Feb | Tianjin, Shijiazhuang, Suzhou, Pingxiang, Jiujiang, Xinyu, Yingtan, Ganzhou, Ji'an, Yichun, Fuzhou, Shangrao, Neijiang, Yibin, Xinyang |
| 7-Feb | Suzhou, Guangzhou |
| 8-Feb | Shenzhen, Foshan, Fangchenggang, |
| 9-Feb | Cangzhou, Huaibei |
| 10-Feb | Beijing, Shanghai |
| 13-Feb | Hohhot, Baotou, Wuhai, Chifeng, Tongliao, Ordos, Hulun Buir, Bayan Nur, Ulanqab, Xing'an League, Xilingol League, Alxa League |

